# SCALING UP THE TASK-SHARING OF AN EVIDENCE-BASED PSYCHOLOGICAL TREATMENT FOR DEPRESSION IN RURAL INDIA

**DOI:** 10.1101/2024.10.23.24315962

**Authors:** Ravindra Agrawal, Mohit Sood, Anushka Patel, Tanushri Sharma, Harshita Yadav, Jigyasa Kaur, Smita Kumari, Prashant Sharma, Vandana Shukla, Balkrishan Tripathi, Namdeo Dongare, Nityasri Sankha Narasimhamurti, Anant Bhan, Sharad Tiwari, Shailesh Sakalle, Vikram Patel

## Abstract

**Background:** Majority evidence on task-sharing of psychological treatments for depression is focused on randomized controlled trials with project staff delivered treatment. Ours is a scaling up of a brief evidence-based psychological treatment (the Healthy Activity Program, HAP) by community health workers (ASHA) in rural India. Our objective was to test the acceptability, feasibility and effectiveness of ASHA delivered HAP.

**Method:** ASHA were recruited in three rural districts in Madhya Pradesh, India. During the study duration, 1001 ASHA completed training using the EMPOWER approach (digital curricula and supervision protocols); 458 ASHA went on to deliver the HAP to adults with depression screened opportunistically. This paper describes the delivery of the HAP over a one-year period (24-07-2022 till 30-06-2023). The primary outcomes were treatment completion, patient and ASHA satisfaction, and change in depression symptom scores on treatment completion; we also explored if treatment effects were sustained at long-term (i.e., 9 month) among a consecutively recruited sub-sample of 10% of the total participants (n=246).

**Results:** 94.3% of the NSPs completed the full training. 2208 patients (12.1% of the total screened) had depression and all 2208 (100%) agreed to receive the treatment. A total of 13,008 sessions were delivered with a 97.82% completion rate. We found substantial reduction in depressive symptom severity from baseline to immediate post-treatment [Cohen’s d=2.52; CI: 2.44 to 2.61], which was sustained at 9-month follow-up [Cohen’s d=.96, 95% CI: .81 to 1.11]. Lower baseline depression, male gender, longer treatment duration, and higher educational status of the ASHA predicted better treatment outcome at endline. Both ASHA and patients reported high levels of satisfaction.

**Conclusion:** The scaling up of a brief evidence based psychological treatment by existing frontline workers through digital platforms for training and supervision is associated with both high levels of satisfaction, treatment completion and remission rates.

## INTRODUCTION

Task sharing of the delivery of psychosocial interventions by front-line non-specialist providers has emerged as an evidence-based approach to address the vast unmet needs for mental health care, in particular in low-resource settings. Scores of randomized controlled trials from around the world, including high-income countries, have demonstrated the effectiveness of task sharing of psychosocial interventions for the prevention and care of a range of mental health conditions(1–3). These interventions typically incorporate a single or a few “active ingredients” and can be delivered in a few sessions in community or primary care settings(4), or remotely, by using digital technology(5).

One such intervention is the Healthy Activity Program (HAP), a behavioral activation-based psychological treatment for depression. The HAP is delivered over 6-8 sessions and leads to high rates of remission and recovery over 12 months(6,7). The effectiveness of the HAP has been replicated in Nepal(8) and the intervention has been adapted for perinatal depression in North America (9). Despite behavioral activation being recommended as a first line treatment for depression (10), the vast majority of persons living with this condition do not have access largely due to shortage of skilled providers and demand side barriers related to stigma attached to seeking mental health care. The scaling up of interventions like the HAP is a priority for global mental health to improve population-wide access after treatments have been found effective in and across similar settings.

In order to make this psychological treatment available at scale, tens of thousands of frontline providers will have to be trained and supervised. Therefore, the conventional face-to-face, expert-led, training and supervision models which dominate the methodology of randomized controlled trials, presents a formidable barrier to scaling up treatment training and delivery. EMPOWER (www.empowerindia.care) is a Harvard-Sangath initiative which deploys a range of digital tools, methods and procedures to rapidly build the capacity of a frontline workforce to deliver evidence-based psychosocial interventions. The program involves selection of evidence-based interventions, development of competency-based digital learning content followed by digitally delivered supervision, assessment of competencies and continuing quality assurance(11).

In this study, we describe the first effort to scale up the task-sharing of any psychological treatment in India, and possibly one of a handful of similar efforts from any low-resource country, which has deployed the EMPOWER approach train frontline workers of India’s National Health Mission (the ASHA) to deliver the HAP to their communities in three rural districts of Madhya Pradesh. The objectives of this study are to evaluate the acceptability and feasibility of the EMPOWER approach to scale up HAP through delivery by ASHA and the effectiveness of the HAP in promoting remission (at the end of treatment) and recovery (sustained remission 9 months after treatment completion). We also sought to identify patient, provider, and treatment-related predictors of depression at each timepoint.

## METHOD

### Partners and Setting

This study was embedded in a large-scale implementation project to address unmet needs for care for depression in rural communities in three districts of Madhya Pradesh, a central India state with a population of about 72 million. The state is one of the least resourced states in India. The scarcity of trained mental health professionals and their concentration in mental health institutions located in urban areas contribute to a treatment gap of 91% for all mental health problems(12). The project was implemented by Sangath, a NGO with a long history of mental health implementation science and which had developed the original HAP intervention, in collaboration with the National Health Mission and the Directorate of Health Services of the Government of Madhya Pradesh. Spanning a 2-year period (2021–2023), the project involved engaging with the primary health system in these rural districts, obtaining necessary permissions, training the ASHAs, screening and delivery of the HAP, independent outcome assessments for a sub-sample of patients, and public engagement events.

### Ethics Approval

The research was approved by Sangath Institutional Review Bpard [protocol number: RA_2023_90].

### Stakeholder/Gatekeeper engagement

Concurrently to the training of ASHAs, we designed a digital course on ‘Leadership in community mental health’ with the aim of building capacity of health system stakeholders. This course contained lectures from knowledge leaders on community mental health as well as ‘case stories’ about other community mental health projects across the country. This course was intended to secure buy-in and support from the health system for our on-ground activities.

We also recruited volunteers from the community and trained them to conduct screening for depression in the community. These volunteers included village elders, panchayat (village governing council) members, teachers, social workers, and college students. They also participated in and contributed to our public engagement activities to encourage a demand for depression care in the community.

Lastly, we also engaged with Community Health Officers (CHO), a mid-level health system cadre situated at the Health and Wellness Centers (HWCs)(13). An HWC is the most distal unit of primary care in India and typically caters to a population of 5000 individuals in Madhya Pradesh. The CHOs were trained in the WHOs mhGAP program and supported the ASHAs whenever they encountered a severely ill patient or who did not improve despite having received the full dose of HAP. We trained 214 CHOs across the three districts where this project was implemented. The CHOs in turn were able to access the specialist advice via the telemedicine infrastructure established under the *Ayushman Bharat* scheme. In collaboration with the District health system, we established a referral and support pathways (see Figure 1).

**Figure 1.**
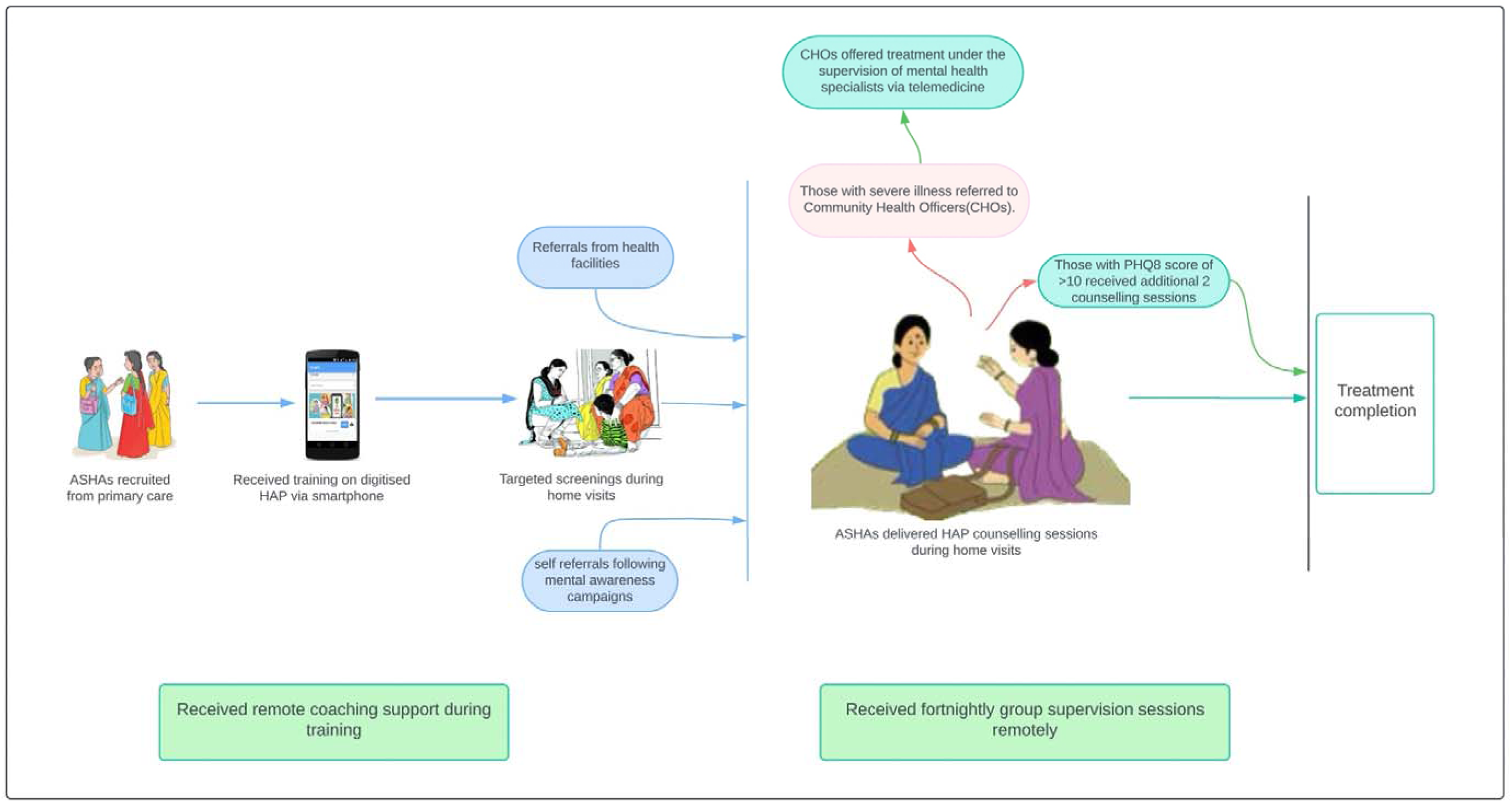
Psychosocial Treatment of Depression in the Community: Referral and Support pathways. ASHAs (Accredited Social Health Activists) were trained in the Healthy Activity Program (HAP) via a smartphone-based Learning Management System (LMS) with ongoing coaching from the project team. Patients were recruited through targeted screenings, referrals, or self-referrals after awareness campaigns. ASHAs provided home-based counseling, receiving supervision every two weeks. Patients with severe conditions were referred to Community Health Officers (CHOs) for treatment under mental health specialists’ supervision. Those with a PHQ-8 score above 10 after initial sessions were given two additional counseling sessions.

### Providers

ASHAs (Accredited Social Health Activists) are India’s most numerous (nearly 1 million) and widely distributed frontline workers. ASHAs, who are all women by design, are incentive based cadres whose original mandate was to promote maternal and child health in rural communities. With the improvements in these health indicators, thanks in large measure due to their deployment, their roles have expanded to addressing issues related to noncommunicable diseases and, most recently, to the door-to-door COVID-19 vaccination campaign. Each ASHA caters to about 500 households, is embedded in the community, reports to the Block Medical officer and works in close collaboration with the Community Health officer (CHO) in which her community is located, and enjoys trust of and access to people’s homes whom she regularly visits. ASHA routinely visit their assigned households to provide a range of health services, actively engage with the communities and are well versed with the health needs of the households where they serve as a ‘last mile’ health worker. ASHAs are affectionately called ‘Didi’ which means elder sister by the community members.

We recruited ASHAs through purposive sampling (based on their availability, access to a smartphone and digital literacy). They were required to use their phones to complete a fully digitized course to learn the HAP. This digitised course had previously been tested(14) in a nearby district; two trials observed that the effects of digitised training with coaching support on provider competencies was non-inferior to the gold standard of expert-led in-person training(14,15). We conducted a 4-hour orientation session in local health facilities in which the project team guided the ASHAs to download the Learning Management System and navigate its features. Subsequently, ASHAs were required to follow the digital curriculum to learn the HAP. The project team monitored their progress and offered remote coaching for trouble-shooting and address queries. As the learner progressed through each module, she undertook an end-of-module assessment. On completion of all the modules, a ‘course completion certificate’ was triggered by the LMS. Following the digital training, ASHAs were required to complete an internship phase involving delivering treatment under supervision by HAP trained counselors of Sangath team to a minimum of two patients.

We provided the ASHAs with a casebook, which contained prompts on session content to be delivered during each visit as part of the structured HAP counselling package. As they delivered counselling sessions, they received fortnightly group supervision sessions and refresher trainings in which they participated in role plays to further reinforce their counselling skills. The supervision sessions were conducted remotely via zoom calls, with each supervision session bringing together 6-8 ASHAs. Supervision sessions were facilitated by trained HAP supervisors who had either a postgraduate qualification in psychology or in social work. During every supervision session, one of the ASHAs presented a session of one of her patients and all group members then discussed the case and gave feedback. As some of the ASHAs gained experience and skills, they were invited to lead the supervision sessions in line with the evidence-based peer supervision strategy for scale-up and sustainability(16,17). ASHAs received incentives for undertaking training, conducting counselling sessions and attending supervision sessions as per the local health system rates.

### Patient recruitment

We administered the Hindi version of PHQ-8(18) to identify patients; while the full version of the PHQ has nine items, the ninth item, referring to suicidal ideation, was removed systematically because local experience demonstrated that this item was rarely endorsed and led to discomfort for both the ASHA and the respondent. Even so, the PHQ-8 is a widely used measure for assessing symptoms of depression and has good sensitivity and specificity(19); further, the 8 item version has been validated among Hindi-speaking populations(20).

ASHAs were encouraged to screen adults during their routine household visits. Individuals scoring 10 or above were considered to have clinically significant depressive symptoms and were offered the 6-session HAP. After completion of the treatment, the PHQ-8 was reassessed, and those still scoring 10 or above were provided with 2 additional counseling sessions. If high scores persisted after the 8th session, referral to a community health officer (CHO) was initiated, with treatment overseen by a mental health specialist (Figure 1).

### Patient and Public Involvement

Patients played a crucial role in the planning and implementation of our project, significantly influencing the delivery of psychological treatment. Patients’ feedback on issues related to (a) Stigmatization i.e. apprehension about receiving mental health-related services in public health facilities and (b) accessibility issues of the health facilities being located at considerable distances, posing challenges, particularly for women, prompted to adopt a visit-at-home strategy for treatment delivery. At times, patients suggested alternative locations such as nearby temples, schools, or panchayats, due to discomfort in discussing their confidential issues in presence of other family members.

Community members actively participated in dissemination events designed to raise mental health awareness. These events included street plays and a reel-making competition, both of which were well-received and effectively engaged the community in mental health discourse.

### Data Collection

We collected a range of data:

#### ASHA training and supervision session attendance

We generated back-end data from the LMS to estimate the number of days taken to complete the online training. Attendance logs for supervision were maintained by the project team for all the ASHAs who were delivering sessions.

#### Patient treatment engagement and outcome scores

We collected process data on screening numbers, patient recruitment, baseline and end-line PHQ-8 scores, counselling session counts, duration of treatment. Nine-month PHQ-8 scores were collected through independent assessors by deploying consecutive sampling to achieve a subsample size of 10% of the original cohort. Outcome data could not be collected for treatment dropouts (*n*=48, 2.17%).

#### Satisfaction questionnaires

We administered satisfaction questionnaires to a subsample of ASHAs and patients. The ASHA questionnaire comprising 6 items rated on a five-point Likert scale was administered to a subset of the providers (*n*=256). The patient questionnaire comprising 5 items was administered to a subset of the patients (*n*=755).

### Analyses

All analyses were conducted on STATA version 17.

We did a descriptive analysis for analyzing socio-demographic ASHA (education, age, marital status) and patient (age) data. Descriptive analysis was also deployed to analyze the project implementation data of ASHA performance metrics (days taken to complete HAP course, number of patients seen) and therapy metrics (Baseline PHQ-8, Post treatment PHQ-8, Independent PHQ-8, treatment duration).

Responses to ASHA and patient satisfaction questionnaire were analyzed descriptively. A two-level mixed-effect regression model was employed to examine the relationship between various factors - including baseline PHQ-8 score, patient age, gender, treatment duration, ASHA age, and ASHA education - and the endline PHQ-8 score. This model addressed the clustering of data by ASHAs, capturing both correlated effects and variations between different providers. The Wald chi-square test detected a significant effect (χ² = 30.43, p < 0.001), highlighting the importance of addressing variability at the group level. Our model, which achieved a log likelihood of -3811.788, demonstrated superior fit compared to alternative approaches.

## RESULTS

### ASHA and Patient characteristics (Table 1)

We received permission from the health system to train 1061 ASHAs. Of these, 1001 ASHAs completed the EMPOWER training. The average duration to complete the training was 43.8 days (95% CI, 40.66, 47.04). Due to health system and budgetary constraints and ASHAs’ existing workload, only 458 of the trained ASHAs engaged with the delivery of the HAP in their catchment areas. During the study period, these 458 ASHAs conducted community screenings of depression with 18195 individuals among whom 2208 (12.1%) screened positive and were offered HAP therapy. (Figure 2) The socio-demographic characteristics of providers and patients is described in Table 1. All ASHAs were women with a mean age of 35.88 years with 95%CI [35.33, 36.43]. The majority of patients were female (71.3%; n=1540) with mean age of 38.92 years with 95% CI [38.42,39.43].

**Figure 2.**
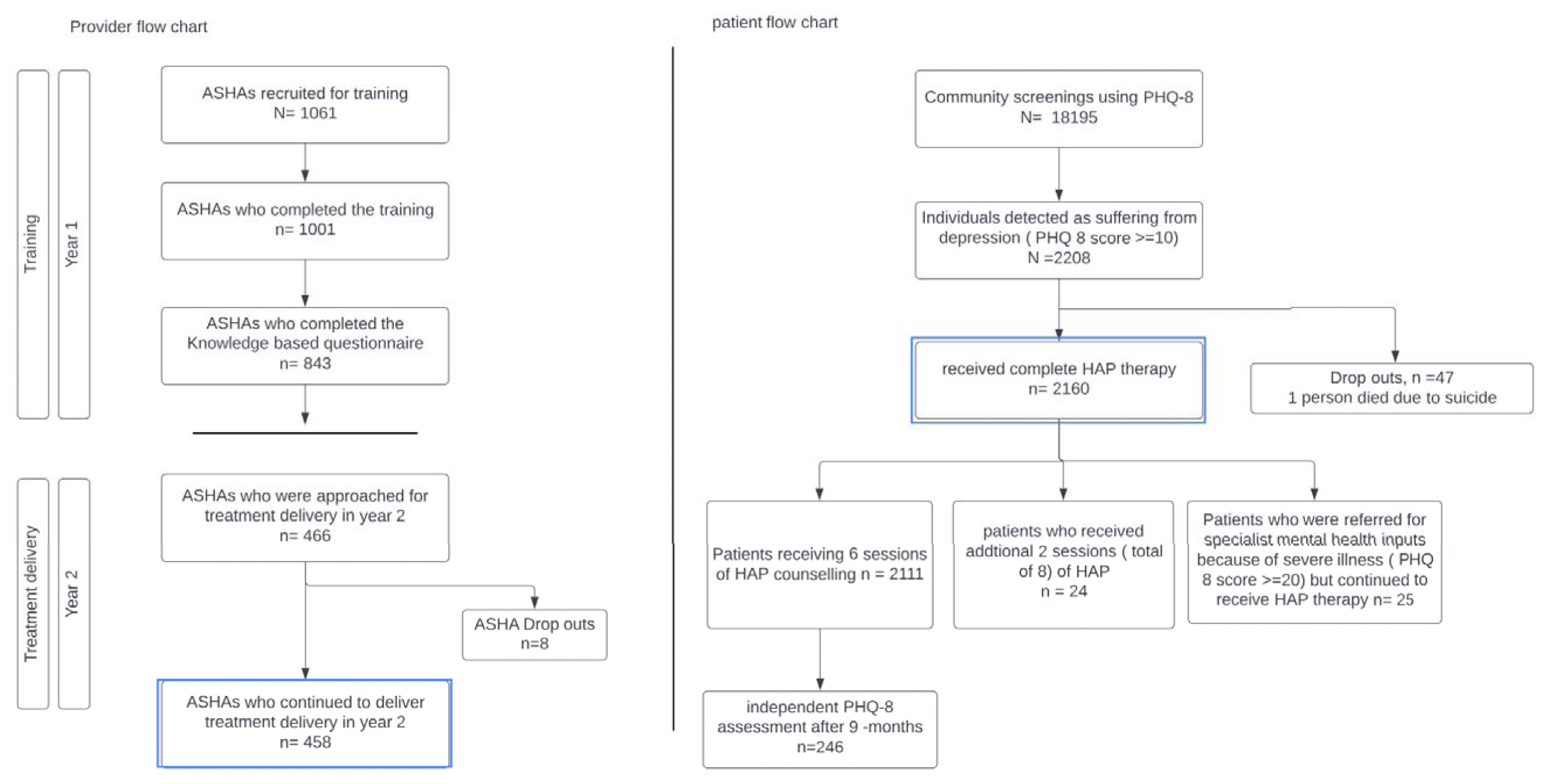
– Provider and patient journey Of the 1061 ASHAs who were recruited for training, 843 completed the post training knowledge assessment. Due to factors related to permissions and workload, 466 were available to deliver treatment of which 8 dropped out. These ASHAs conducted 18195 screenings with 2208 patients being positive for depression and offered treatment. 216o patients completed HAP therapy with about 47 dropouts and 1 death due to suicide.

**Table 1.**
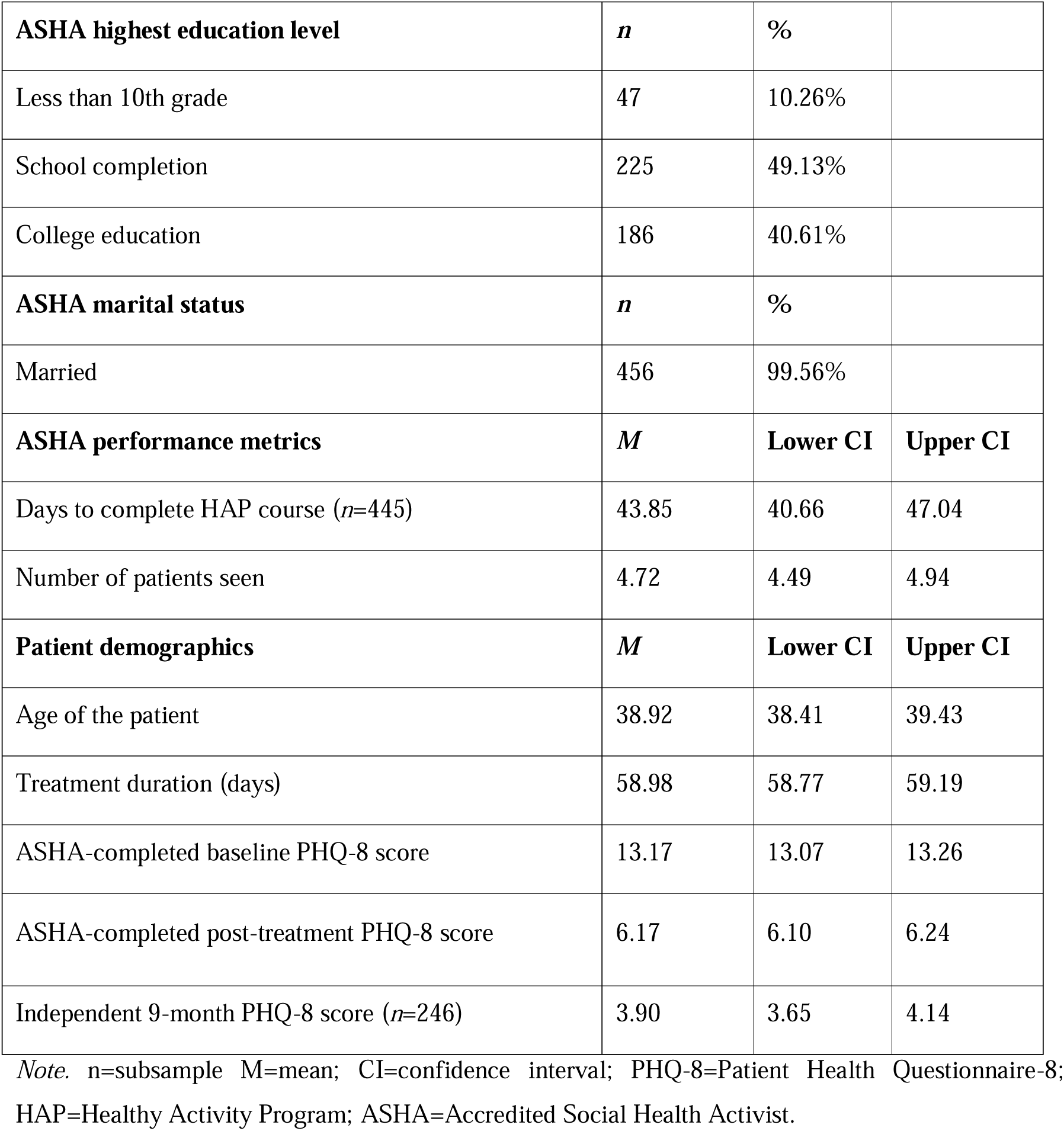
Provider (ASHA) (*N*=458) and Patient (*N*=2160) sociodemographic characteristics.

### Implementation

In the sample of those who scored >9 on the PHQ, 75% had moderate depression (PHQ-8 score 10-14) and the remainder has moderately severe depression (PHQ-8 score 15-19); 25 persons with severe depression were referred to a mental health specialist while also continuing to receive HAP from the ASHAs. There was one death due to suicide – this patient was detected as having severe depression; however, this patient died by suicide before any formal therapeutic help could be organized. 42 patients were referred by the volunteers and 6 by the health facilities; the remainder were screened by the ASHA. Of the 2208 patients who started therapy, 48 patients dropped out; 2160 received 6 counselling sessions, with 24 (1.08%) needing an additional 2 sessions of counselling as per the HAP protocol. A total of 13,008 counselling sessions were delivered and the average duration needed to complete the treatment was about two months (mean 59 days). Nearly all of the sessions were conducted in patients’ homes; less frequently used sites were the local panchayats, schools and places of worship. 452 ASHAs engaged with group supervision conducted on a fortnightly basis.

### Acceptability of the ASHA delivered HAP (Figure 3)

Overall, the ASHAs reported high scores of satisfaction level (>4) for all individual items implying high levels of acceptability with the HAP training and delivering HAP counselling. Similarly, patients reported a high satisfaction score (>4) for all individual items implying higher level of acceptability of receiving HAP therapy.

**Figure 3.**
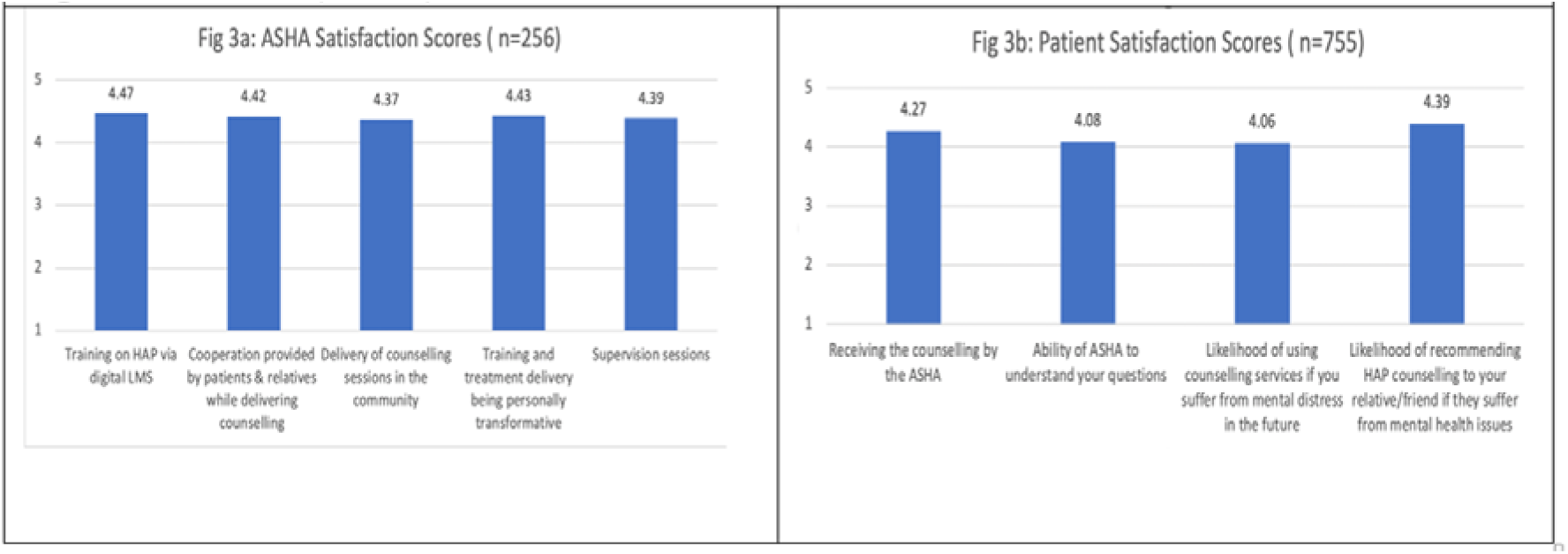
– Provider and patient satisfaction scores. ASHAs expressed high satisfaction (>4) with both the HAP training and their experience delivering counseling. Patients also reported positive feedback, with high ratings (>4) for receiving HAP therapy. This shows that the program was well-received by both those delivering and those receiving care.

### Effectiveness of the HAP delivery

The mean PHQ-8 score reduced from 13.17 to 6.17, representing a reduction of about 50% from the baseline (*t* (2159) = 117.28, *p* < .001]. This translates to a large effect size [Cohen’s *d* = 2.52; CI = 2.44 to 2.61]. These findings were replicated for the independent follow up at 9 months after treatment (*n*=246) at which time we observed a sustained reduction in depressive symptom severity [*t*(248) = 15.21, *p* < .001] which also translates to a large effect size [Cohen’s *d* = .96, 95% CI: .81 to 1.11]. Baseline PHQ-8 score (β =0.037, p<0.05) and treatment duration (β =0.029, p<0.001) were associated with the endline PHQ-8 score (Table 2)

**Table 2.** Predictors of depression severity at the end of treatment (N=2160) (Results of two-level mixed-effect regression model)

| **Predictors** | **Coefficient ($\beta$)** | **Standard error** | **z value** | **p value** | **95% Confidence Interval** |
| --- | --- | --- | --- | --- | --- |
| Baseline PHQ-8 score | 0.037 | 0.015 | 2.440 | 0.014 | [0.007,0.066] |
| Patient age | 0.005 | 0.002 | 1.900 | 0.057 | [0.000,0.010] |
| Patient gender | -0.006 | 0.064 | -0.090 | 0.931 | [-0.131,0.120] |
| Treatment duration | 0.029 | 0.007 | 4.260 | 0.000 | [0.016,0.042] |
| ASHA education | -0.052 | 0.055 | -0.950 | 0.340 | [-0.160,0.055] |
| ASHA age | 0.001 | 0.011 | 0.140 | 0.891 | [-0.020,0.023] |
| Intercept | 3.926 | 0.620 | 6.330 | 0.000 | [2.711,5.141] |
| **Random effects** |  |  |  |  |  |
| **Group** | **Estimate** | **Standard error** | **95% Confidence Interval** |  |  |
| Intercept (ASHAs) | 1.548 | 0.128 | [1.317,1.821] |  |  |
| Residual | 1.378 | 0.047 | [1.288,1.474] |  |  |

## DISCUSSION

This paper describes the experience of the scaling up an evidence based psychological treatment by existing community health workers in rural populations in India. The scaling up was catalyzed by using a digital platform that is accessible via smartphones, to enable large-scale training of ASHAs. This approach was remote coach-supported, allowed ASHAs to complete the training in a self-paced manner, and attend supervision sessions remotely. We observed high feasibility and acceptability as evidenced by a range of provider and patient metrics including: high levels of completion of the digital training; high levels of patient engagement and completion of the ASHA delivered multi-session treatment; and high levels of provider and patient satisfaction with the program. Overall, we observed large improvements in depression severity at the end of treatment. We found that lower baseline depression severity and longer treatment duration predicted lower depression scores at treatment completion. Moreover, we found a sustained effect on patient depressive scores nine months later. Patient age and gender and the ASHA age and education levels were not associated with the intervention effectiveness indicating the applicability of the delivery to a diverse patient group and by a wide range of community-based providers.

This study builds upon the large, randomized controlled trial, evidence base on the effectiveness of front-line worker delivered mental health care (3,4,21) by providing real-world evidence on the scaling up of such interventions through deploying existing government health providers and a digital platform for training and supervision. There are many barriers to addressing the unmet needs for care for depression in India(22). These include both supply side barriers (notably the lack of skilled providers) but also, importantly, demand-side barriers such as lack of awareness, stigma attached to mental health care, delay in identification and cost of seeking treatment(23–25). Our program addressed both these sets of barriers and capitalized on the fact that the ASHAs shared the same lived experiences as those of individuals who were receiving treatment and enjoyed deep historic relationships with their community members. Their existing health promotion roles allowed for seamless integration of the session delivery schedule into their routine community errands, effectively relieving patients from the burden of travel or needing to take time off work. The strategy of administered treatment through home visits significantly contributed to the achievement of high completion rates.

The effectiveness of the HAP offsets concerns that the original trial findings testifying to its efficacy in controlled trial settings(7) might not generalize in a real-world scale-up. We attribute the implementation success of this project to the high treatment completion rates fostered by the existing rapport, trust, and interpersonal skills between ASHAs and their communities. Moreover, our findings underscore the enduring effectiveness of the intervention, as patients continued to exhibit relief from depression nine months after the conclusion of therapy, demonstrating the sustained utilization of activation skills acquired during treatment(26). Notably, the active engagement of key stakeholders in the overall program design and implementation also played a role in ensuring support for the ASHA to play these new roles and to enable that referral pathways for the small number of patients who needed more specialized care.

Our study had two significant constraints. Firstly, the inability to independently assess sustained outcomes for all participants due to limited resources, as the project was not initially intended as a research study. However, our subsample (*n*=246) was identical with respect to baseline variables, and therefore representative, of the full sample. Second, we did not use the full nine item version of the PHQ due to community level feedback about the low acceptability of this item. That said, the PHQ-8 has very high concordance with the PHQ-9 in screening for depression(19) and we do not expect that the omission of this one item will materially impact our findings(27).

This study demonstrates that it is feasible to use the EMPOWER approach of deploying digital platforms to train and supervise frontline workers at scale to deliver psychosocial interventions and that mental health care by these frontline workers was highly acceptable to themselves and their communities and associated with large clinical benefits. We hope that our findings can inform the strategic planning of the health system to make quality mental health care available and accessible in rural communities. The ASHA being a relatively well-established cadre was a natural choice for delivering the intervention; however, future efforts may also expand this provider base to include other frontline workers in the health system, notably the recently minted Community Health Officers (CHO) who, unlike the ASHA, are full-time employees of the health system. Although our study demonstrates that community treatment of depression via existing front-line workers can be delivered effectively at scale, we note that the project paid the equivalent incentive fee to the ASHA and that sustained scale-up will need adequate budgetary allocation to compensate them to deliver counselling sessions, and support supervision and referrals systems.

## Data Availability

All data produced in the present work are analyzed and are presented in the manuscript.

## Acknowledgements

We acknowledge the efforts of the HAP supervisors (Sweta Dubey, Ritesh Dubey, Gaurishankar Dwivedi, Ramkumar Sapre, Brajesh Prashar, Prateeksha Sharma, Pragya Sharma, Mahendrasingh Kushwah) and all the volunteers. We acknowledge Dr Chirag Patel and Dr Vivek Yadav, DMHP psychiatrists and Dr. Sarjan Singh Sengar (Medical Officer In charge-Mental health) who helped with the CHO training and provided specialist support when needed. We would also express our gratitude to Dr Sharad Tiwari, Dr. Dinesh Khatri (CMHO Raisen), Dr. Dinesh Dahlwar (CMHO Narmadapuram) 2, Dr. Ajay Kumar Upadhyay (CMHO Vidisha) and all the BMOs in the three districts who extended enthusiastic support and guided us throughout the project duration. Lastly, we acknowledge the study participants ( the ASHAs and the patients) and our collaborators from the National Health Mission, Madhya Pradesh State and National Health Systems Resource Center, Delhi, India.

## Author contributions

Ravindra Agrawal lead the project implementation and was responsible for conceptualizing and drafting the manuscript. Prashant Sharma, Vandana Shukla and Balkrishan Tripathi were responsible for data collection and implementation of the project activities at district sites. Tanushri Sharma, Jigyasa Kaur, Harshita Yadav and Smita Kumari were responsible for ASHA training and supervision, data cleaning, and project implementation. Mohit Sood was responsible for designing the data collection forms, project implementation, data cleaning and conducting data analysis. Anushka Patel was responsible for conducting data analysis and providing critical inputs to the manuscript. Namdeo Dongare was responsible for running the regression analysis and providing critical inputs for data analysis. Vikram Patel mentored the team, provided critical inputs for data analysis and reviewed the manuscript. Anant Bhan was co-lead of the project and reviewed the manuscript. Nityasri Sankha reviewed the manuscript. Sharad Tiwari and Shailesh Sakalle were responsible for supervision of the project processes, guidance to mitigating the implementation challenges and helped embed the program within the health system. All authors have critically reviewed this manuscript and provided consent for publication.

## Funding

This work was supported by a CSR grant by the Johnson and Johnson Foundation, India. The funder had no role in study design, data collection and analysis, decision to publish, or preparation of the manuscript.

## Conflicts of interests

The authors declare no competing interests.

## Notes

### Competing Interest Statement

The authors have declared no competing interest.

### Author Declarations

The research was approved by Sangath Institutional Review Board [protocol number: RA_2023_90].

## References

1. Cuijpers P, Karyotaki E, Reijnders M, Purgato M, Barbui C. Psychotherapies for depression in low- and middle-income countries: a meta-analysis. World Psychiatry. 2018;17(1):90–101.

2. Patel V. Scale up task-sharing of psychological therapies. Lancet (London, England) [Internet]. 2022 Jan 22;399(10322):343–5. Available from: http://www.ncbi.nlm.nih.gov/pubmed/34929197

3. van Ginneken N, Chin WY, Lim YC, Ussif A, Singh R, Shahmalak U, et al. Primary-level worker interventions for the care of people living with mental disorders and distress in low- and middle-income countries. Cochrane Database Syst Rev [Internet]. 2021 Aug 5;2021(8). Available from: http://doi.wiley.com/10.1002/14651858.CD009149.pub3

4. Singla DR, Kohrt BA, Murray LK, Anand A, Chorpita BF, Patel V. Psychological Treatments for the World: Lessons from Low- and Middle-Income Countries. 2017;49:149–81.

5. Stein DJ, Naslund JA BJ. COVID-19 and the global acceleration of digital psychiatry. The Lancet Psychiatry. 2022;9(1):8–9.

6. Weobong B, Weiss HA, McDaid D, Singla DR, Hollon SD, Nadkarni A, et al. Sustained effectiveness and cost-effectiveness of the Healthy Activity Programme, a brief psychological treatment for depression delivered by lay counsellors in primary care: 12-month follow-up of a randomised controlled trial. PLoS Med. 2017;14(9):0–47.

7. Patel V, Weobong B, Weiss HA, Anand A, Bhat B, Katti B, et al. The Healthy Activity Program (HAP), a lay counsellor-delivered brief psychological treatment for severe depression, in primary care in India: a randomised controlled trial. Lancet [Internet]. 2017 Jan 14;389(10065):176–85. Available from: 10.1016/S0140-6736(16)31589-6

8. Jordans MJD, Luitel NP, Garman E, Kohrt BA, Rathod SD, Shrestha P, et al. Effectiveness of psychological treatments for depression and alcohol use disorder delivered by community-based counsellors: Two pragmatic randomised controlled trials within primary healthcare in Nepal. Br J Psychiatry. 2019;215(2):485–93.

9. Singla DR, Lawson A, Kohrt BA, Jung JW, Meng Z, Ratjen C, Zahedi N, Dennis CL P V. Implementation and effectiveness of nonspecialist-delivered interventions for perinatal mental health in high-income countries: a systematic review and meta-analysis. JAMA psychiatry. 2021;78(5):498–509.

10. WHO. Depressive Disorder (Depression). https://www.who.int/news-room/fact-sheets/detail/depression. 2023. Accessed on 28/02/2024.

11. Patel V, Naslund JA, Wood S, Patel A, Chauvin JJ, Agrawal R, et al. EMPOWER: Toward the Global Dissemination of Psychosocial Interventions. Focus (Madison). 2022;20(3):301–6.

12. Kokane A, Pakhare A, Gururaj G, Varghese M, Benegal V, Rao GN, et al. Mental health issues in madhya pradesh: Insights from national mental health survey of india 2016. Healthc. 2019;7(2):1–12.

13. Grewal H, Sharma P, Dhillon G, Munjal RS, Verma RK, Kashyap R. Universal Health Care System in India: An In-Depth Examination of the Ayushman Bharat Initiative. Cureus. 2023;15(6):1–5.

14. Muke SS, Tugnawat D, Joshi U, Anand A, Khan A, Shrivastava R, et al. Digital training for non-specialist health workers to deliver a brief psychological treatment for depression in primary care in India: Findings from a randomized pilot study. Int J Environ Res Public Health. 2020;17(17):1–22.

15. Naslund JA, Tugnawat D, Anand A, Cooper Z, Dimidjian S, Fairburn CG, et al. Digital training for non-specialist health workers to deliver a brief psychological treatment for depression in India: Protocol for a three-arm randomized controlled trial. Contemp Clin Trials [Internet]. 2021;102(October 2020):106267. Available from: 10.1016/j.cct.2021.106267

16. Singla DR, Weobong B, Nadkarni A, Chowdhary N, Shinde S, Anand A, et al. Improving the scalability of psychological treatments in developing countries: An evaluation of peer-led therapy quality assessment in Goa, India. Behav Res Ther. 2014;60:53–9.

17. Singla DR, Savel KA, Magidson JF, Vigod SN DC. The Role of Peer Providers to Scale Up Psychological Treatments for Perinatal Populations Worldwide. Curr Psychiatry Rep. 2023;25(11):735–40.

18. Kroenke K, Strine TW, Spitzer RL, Williams JBW, Berry JT, Mokdad AH. The PHQ-8 as a measure of current depression in the general population. J Affect Disord. 2009;114(1–3):163– 73.

19. Kroenke K, Spitzer RL, Williams JBW, Löwe B. The Patient Health Questionnaire Somatic, Anxiety, and Depressive Symptom Scales: A systematic review. Gen Hosp Psychiatry. 2010;32(4):345–59.

20. Patel AR, Prabhu S, Sciarrino NA, Presseau C, Smith NB, Rozek DC. Gender-Based Violence and Suicidal Ideation Among Indian Women From Slums: An Examination of Direct and Indirect Effects of Depression, Anxiety, and PTSD Symptoms. Psychol Trauma Theory, Res Pract Policy. 2021;13(6):694–702.

21. Singla DR, Meltzer-Brody SE, Silver RK, Vigod SN, Kim JJ, La Porte LM, et al. Scaling Up Maternal Mental healthcare by Increasing access to Treatment (SUMMIT) through non-specialist providers and telemedicine: a study protocol for a non-inferiority randomized controlled trial. Trials. 2021;22(1):1–15.

22. Roberts T, Shidhaye R, Patel V, Rathod SD. Health care use and treatment-seeking for depression symptoms in rural India: An exploratory cross-sectional analysis. BMC Health Serv Res. 2020;20(1):1–13.

23. Jani A, Ravishankar S, Kumar N, Vimitha J, Shah S, Pari A, et al. Factors influencing care-seeking behaviour for mental illness in India: A situational analysis in Tamil Nadu. J Public Heal (United Kingdom). 2021;43:II10–6.

24. Roberts T, Miguel Esponda G, Krupchanka D, Shidhaye R, Patel V, Rathod S. Factors associated with health service utilisation for common mental disorders: A systematic review. BMC Psychiatry. 2018;18(1).

25. Lakshmana G, Sangeetha V P V. Community perception of accessibility and barriers to utilizing mental health services. J Educ Health Promot. 2022;11(1):56.

26. Bhat B, De Quidt J, Haushofer J, Patel VH, Rao G, Schilbach F VP. The long-run effects of psychotherapy on depression, beliefs, and economic outcomes. National Bureau of Economic Research.

27. Shin C, Lee SH, Han KM, Yoon HK, Han C. Comparison of the usefulness of the PHQ-8 and PHQ-9 for screening for major depressive disorder: Analysis of psychiatric outpatient data. Psychiatry Investig. 2019;16(4):300–5.

